# Causal factors in primary open angle glaucoma: a phenome-wide Mendelian randomisation study

**DOI:** 10.1101/2022.10.30.22281718

**Authors:** Thomas H Julian, Zain Girach, Eleanor Sanderson, Hui Guo, Jonathan Yu, Johnathan Cooper-Knock, Graeme C. Black, Panagiotis I Sergouniotis

## Abstract

Primary open angle glaucoma (POAG) is a chronic, adult-onset optic neuropathy associated with characteristic optic disc and/or visual field changes. With a view to identifying modifiable risk factors for this debilitating condition, we performed a ‘phenome-wide’ univariable Mendelian randomisation (MR) study and analysed the relationship between 9,661 traits and POAG. Data were analysed using the weighted median method, weighted mode based estimation, the Mendelian randomisation (MR) Egger method and the inverse variance weighted approach. Our analysis identified 11 traits related to POAG risk including: serum levels of the angiopoietin-1 receptor (OR [odds ratio] = 1.11, IVW [inverse variance weighted] p= 2.34E-06) and the cadherin 5 protein (OR= 1.06, IVW p= 1.31E-06); intraocular pressure (OR=2.46 - 3.79, MRE IVW p=8.94E-44 - 3.00E-27); diabetes (beta=1.64, IVW p = 9.68E-04); and waist circumference (OR = 0.79, IVW p=1.66E-05). Future research focussing on the effects of diabetes, waist circumference, serum cadherin 5 and serum angiopoietin-1 receptor on POAG development and progression is expected to provide key insights that might inform the provision of lifestyle modification advice and/or the development of novel therapies.

## INTRODUCTION

Glaucoma is a heterogeneous group of disorders characterised by progressive retinal ganglion cell degeneration and distinctive optic disc changes^1,2^. A key clinical consequence of glaucoma is progressive visual field loss that can affect central vision and lead to irreversible blindness ^3^. The most common glaucoma subtype is primary open angle glaucoma (POAG), a chronic optic neuropathy associated with open iridocorneal angles and an apparent absence of unequivocal secondary causes ^4^. In 2020, the global adult population of POAG sufferers was estimated to stand at 53 million; this is projected to increase to around 80 million affected individuals by 2040 ^5^.

Major risk factors for POAG include increasing age and raised intraocular pressure (IOP) ^6^. Several genetic contributors have also been described including 127 statistically independent single nucleotide variants (SNVs) associated with glaucoma at a genome-wide significant level, indicating potential contributions of several genes to POAG pathogenesis (e.g. *SVEP1, RERE, VCAM1, ZNF638, CLIC5, SLC2A12, YAP1, MXRA5, SMAD6* and *MYO7A*) ^*7,8*^.

Whilst several strategies exist to reduce glaucoma progression, including surgical, laser and topical drug-based approaches, most of these target a single underlying mechanism: raised IOP. Notably, a proportion of individuals with POAG have normal IOP, while a proportion of individuals with high IOP never develop glaucoma. It is therefore widely accepted that additional pathophysiological processes are contributing to the development of POAG. Identifying such IOP-independent mechanisms has the potential to inform complementary therapeutic approaches for this common cause of visual impairment. Further, accruing evidence on the impact of lifestyle factors and interventions on POAG progression is expected to give affected individuals the opportunity to personally impact their prognosis through behavioural change. It is noted that reversing the optic nerve damage that defines glaucoma is presently not possible and, as such, identifying novel strategies to prevent the development and/or to modify the progression of the disease remains of great interest.

Mendelian randomization (MR) is a statistical approach that uses genetic variation to study the causal relationships between modifiable *exposures* (such as body mass index) and medically-relevant *outcomes* (such as risk of a specific disease) ^9,10^. MR offers advantages over conventional epidemiological studies due to the ability of this method to obtain causal effect estimates that are not biased by confounding and reverse causation (which occurs when the outcome precedes and causes the exposure instead of the other way around). In MR, genetic variants in the form of SNVs, are exploited as instrumental variables which can be utilised to study the causal relationship between an exposure variable and an outcome of interest. Its principles are based on Mendel’s laws of segregation and independent assortment, which state that offspring inherit SNVs randomly from their parents and randomly with respect to most other locations in the genome. As a result, genetic variants that are associated with an exposure of interest can be used to proxy the part of the exposure variable that is independent of possible confounding influences (e.g. from the environment and other traits). Further, because genetic variation is necessarily ‘upstream’ of disease and environmental exposures in terms of mechanism, the risk of reverse causation is dramatically reduced ^9^.

To date, the use of MR approaches in the context of POAG has been relatively limited. Despite this, these methods have been successfully implemented to explore the relationship between POAG and a small number of traits including brain alterations ^11^, plasma lipid levels ^12^, fatty acids ^13^, type 2 diabetes ^14^, HbA1c levels ^14^, fasting glucose, coffee consumption ^15^ and blood pressure ^16^.

In this study, we discuss the findings of a systematic, broad (‘phenome-wide’) univariable MR analysis and report a set of traits that show evidence of a causal effect on POAG. Our method pointed to a number of potential novel causal factors and provided further evidence on the role of known glaucoma risk factors.

## RESULTS

Overall, 9,661 traits were eligible for analysis. Of these, 11 were identified to have statistically robust relationships with POAG in a univariable analysis (Table 1 & Supplementary data 1.1). It is noted that 264 traits were related to POAG in a false discovery rate (FDR) corrected inverse variance weighted (IVW) and leave one out analysis; however, there was not agreement about the statistical significance of these signals among all MR methods used (suggesting weaker evidence of a causal effect; supplementary data 1.2). The results for all traits considered are presented in supplementary data 1.3.

**Table 1:**
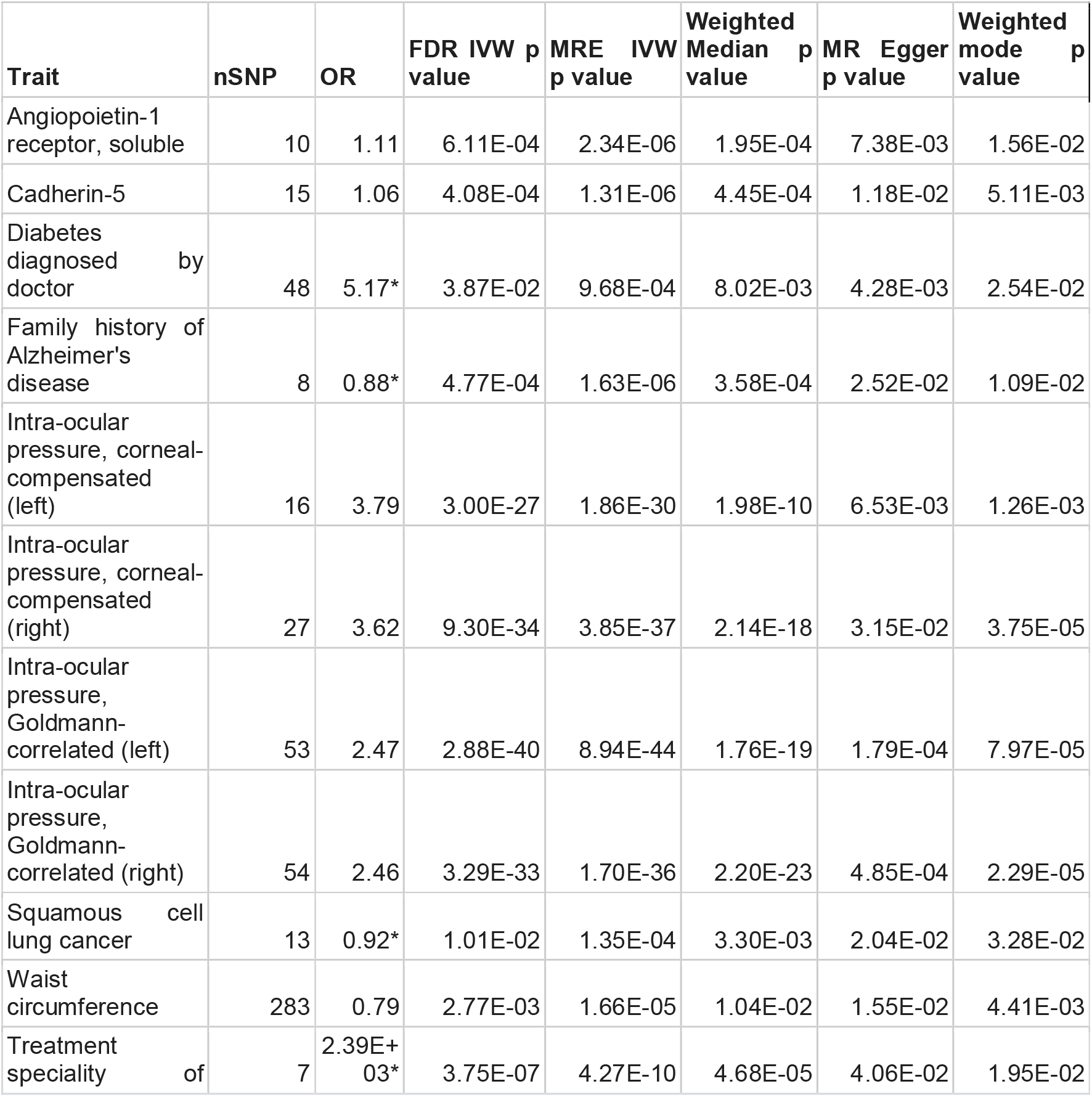
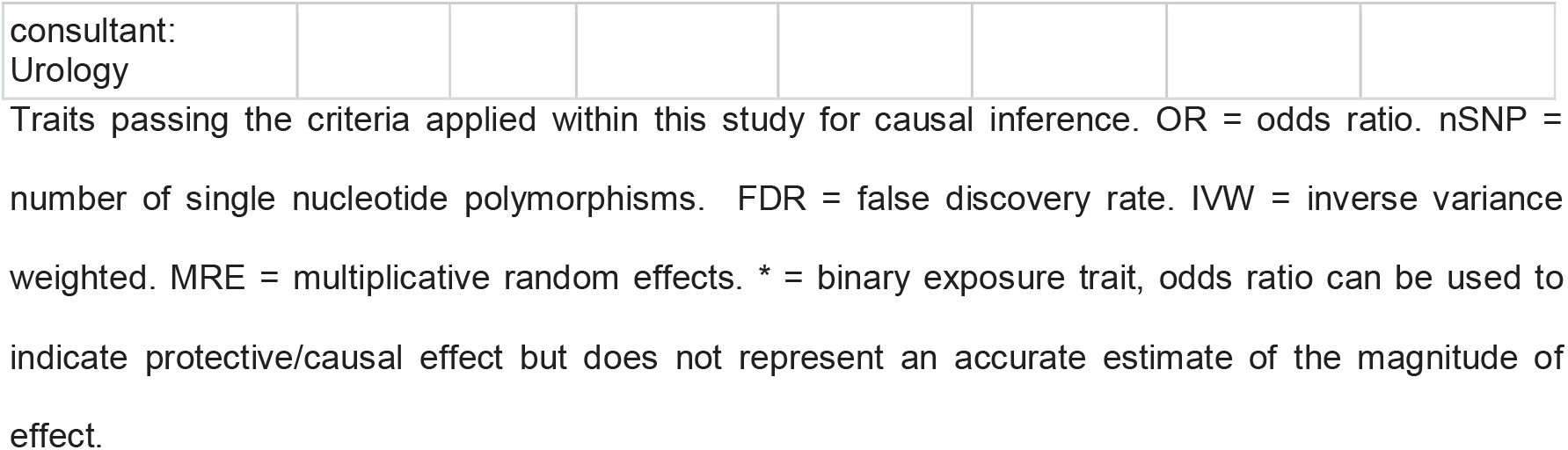
Traits which were linked to POAG risk across robust tests and in an FDR corrected IVW. nSNP = the number of SNPs included in the analysis.

Serving as a positive control, IOP was identified to represent a causal factor for POAG. Both right (OR=2.46, IVW p= 1.70E-36) and left (OR=2.46, IVW p value = 8.94E-44) eye Goldman-correlated measures calculated using a Reichert ocular response analyser (ORA) were demonstrated to be causal for POAG. Additionally, ORA corneal compensated IOPs (that were measured in the same study) were also significantly related to POAG risk (left eye OR=3.79, IVW p value = 1.86E-30; right eye OR=3.62, IVW p = 3.85E-37). This is in keeping with our existing understanding of POAG and validates our methodology.

Waist circumference, a measure of abdominal obesity, was inversely related to POAG risk (OR=0.79, IVW p=1.66E-05, weighted media p=0.01, MR Egger p=.02, weighted mode p=0.004). Whilst body mass index (BMI) was significantly related to POAG risk in IVW (OR=0.84, p=1.93e-05), this did not persist throughout all other MR analyses that we conducted (weighted median p=0.01, weighted mode p=0.13, MR Egger p=0.78) and, as such, the evidence for a protective effect was less strong.

Our screen identified two serum proteins with a causal effect on POAG. These are angiopoietin -1 receptor (OR= 1.11, MRE IVW p= 2.34E-06) and cadherin-5 (OR= 1.06, MRE IVW p= 1.31E-06). Orthogonally we discovered that rare deleterious missense variants within the gene encoding angiopoietin-1 receptor (*TEK*) increase risk for POAG (OR=2.48, p=0.009). In contrast, rare variant burden testing did not reveal a relationship between changes in the gene encoding cadherin-5 (*CDH5*) and POAG ^17^.

Diabetes (diagnosed by a doctor) was identified to be causally related to POAG (MRE IVW beta= 1.64, MRE IVW p= 9.99E-04). Individual diabetes subtypes were also explored. Type 2 diabetes was not found to be causally related to POAG risk (IVW p= 0.21). However when data from a genome-wide association study (GWAS) that adjusted for BMI were used as input, type 2 diabetes was found to be causally related to POAG in two measures (IVW beta=0.03, IVW p=0.003, weighted median p=0.009, MR Egger p=0.10, weighted mode p=0.08) ^18^. Whilst acknowledging the potential significance of this finding, it is important to note that using covariable-adjusted exposure instruments can lead to bias in MR (albeit to a lesser extent than bias created by coviarable-adjusted outcome instruments) ^19^. Type 1 diabetes was nominally significant in IVW analysis in relation to POAG (IVW beta=0.02, p=0.03) but no statistically significant signals were obtained for this trait by the other MR methods used in this study (weighted median p=0.07, weighted mode p=0.10, MR Egger p=0.19).

Our screen identified family history of Alzheimer’s disease as a protective factor for POAG (beta - 0.13, IVW p value=1.63E-06). Given this is a family history trait, it appears probable that the analysis would violate the third key assumption of MR (i.e. that the genetic instruments are likely to be associated with the outcome via horizontal pleiotropy rather than through vertical pleiotropy). Alzheimer’s disease itself was not related to POAG risk (IVW p value=0.96).

Squamous cell lung cancer is suggested to be a protective trait for POAG from our MR results (MRE IVW beta= -0.08, MRE IVW p = 1.35E-04). It is however known that survival bias can distort MR results, and thus this result should be treated with caution ^20,21^.

Intriguingly, individuals under the care of a urology service were identified to have a higher risk of POAG (beta=7.78, IVW p= 4.27E-10). This of course represents a composite trait confounded by urological phenotypes,risk factors and medicine use. It is not possible to discern the nature of these confounding factors within this study.

## DISCUSSION

In this study, we used MR analysis to advance our understanding of POAG pathogenesis. We outline a robust approach to causal inference that can be applied in other disorders with wide-reaching implications. The validity of our methodology is supported by the identification of IOP as the leading causal trait for POAG; highlighting this well-established risk factor essentially serves as a positive control. Notably, our MR analysis supported causal roles for several disputed risk factors and identified novel causal traits.

Our findings support a causal effect between a diagnosis of diabetes and POAG. This is in accordance with an extensive literature base which also supports this relationship ^6^. Furthermore, in patients with existing POAG, diabetes has been shown to advance the severity of glaucoma in the form of raising IOP, worsening visual field defects and resulting in a larger cup-disc ratio ^22^. Several mechanisms have been proposed to account for the relationship between diabetes and POAG. First, it has been suggested that IOP could be raised due increased formation of advanced glycation end products causing trabecular meshwork cell damage and resulting in blockage of aqueous humour outflow through the iridocorneal angle; serum/aqueous hyperosmolarity and hyperviscosity may also increase aqueous humor retention which could also impact IOP ^23^. It has also been proposed that long standing hyperglycaemia and lipid abnormalities might increase the vulnerability of neurons to stress and, as such, increase vulnerability to POAG development ^6^.

There is controversy surrounding the role of adiposity in POAG. There are numerous measures of adiposity including BMI, waist circumference, waist-hip ratio and body fat percentage; each of these measures has its own strengths and weaknesses ^24^. For instance, it is understood that BMI in isolation does not serve to deliver sufficient information regarding adiposity to fully evaluate cardiometabolic disease risk. As a result, it has been recommended that waist circumference is adopted as an additional routine measurement to identify and classify obesity ^24–26^. Although lower BMI has been associated with worse glaucoma outcomes in some studies, this finding is not universal^27^. Further, some studies have identified a causal relationship between obesity and POAG ^28^. Here, we find that waist circumference is inversely associated with POAG risk and, as such, increasing waist circumference is protective for POAG. The evidence for a relationship with BMI is less compelling but it is noted that this measure does not necessarily serve as a good measure of fat distribution, abdominal obesity or fat/fat free mass ^24^. Our findings are at odds with the conclusions of a recent MR study by Lin et al., which described a causal relationship between obesity and POAG ^29^. The principal difference between our analysis and that of Lin *et al*. is our use of a larger POAG population (15,229 cases and 177,473 controls versus 1,824 POAG cases and 93,036 controls). It is also noted that the conclusions by Lin *et al*. are not strongly supported by some of their MR analyses which did not reveal statistically significant signals. This may suggest that their results are vulnerable to outlier SNVs and horizontal pleiotropy. In summary, our findings suggest that abdominal obesity may be associated with decreased risk of POAG and offer competing evidence with the study by Lin *et al*. Given these competing results and the remaining controversy ^26^ further work is warranted in this area.

Cadherins are a group of transmembrane proteins responsible for cell adhesion. These molecules have received little attention in relation to POAG although a small number of studies describe a potential role in this condition. In a previous genome-wide copy number variant study, cadherin signalling was implicated in POAG susceptibility ^30^. The authors of that study noted that cell adhesion proteins determine trabecular meshwork resistance to aqueous outflow and, as such, may impact IOP and therefore have a role in glaucoma. Our MR results have shown a causal effect of cadherin-5 on POAG (although further evidence on this relationship could not be obtained through rare variant burden testing). Cadherin 5 (also known as vascular endothelial (VE) cadherin) is not known to be expressed in trabecular meshwork cells, but it is expressed in the endothelial cells of Schlemm’s canal ^31^. Notably, a recent study by Kelly *et al*. reported that primary Schlemm’s canal cells from individuals with glaucoma had higher levels of cadherin 5 protein expression compared to cells from healthy donor eyes ^32^. It was therefore speculated by the authors that cadherin 5 may increase aqueous outflow resistance through decreasing paracellular permeability. Whilst it is important to note that our study relates to serum cadherin 5 rather than expression local to the anterior segment, our results do support a role for this protein in POAG pathophysiology.

Angiopoietin-1 receptor is a cell surface receptor encoded by the gene *TEK*. Angiopoietin proteins are vascular growth factors involved in the development and remodelling of blood vessels, and angiopoietin-TEK signalling is pivotal to the development of Schlemm’s canal ^33^. Previous work in mice has demonstrated that angiopoietin has a critical role in preserving the integrity of Schlemm’s canal and it has been suggested that abnormalities of angiopoietin physiology may have a role on POAG ^34^. Similarly, human genetic studies have provided support for a role for angiopoietin physiology in POAG, with common variants in the angiopoietin-1 receptor ligand (*ANGPT1*) having been found to influence IOP ^35^ and mutations in *TEK* causing primary congenital glaucoma ^36^. Our MR analysis supports a causal role for serum angiopoietin-1 receptor levels in POAG and our rare variant burden test suggests that loss of function of the corresponding gene (*TEK*) is linked to POAG development. Whilst the MR and rare variant analyses are not concordant with respect to the direction of effect, this may represent differential expression of angiopoietin-1 receptor intraocularly and in the serum. As for cadherin 5, it is outside the scope of this study to investigate the local effects of angiopoietin-1 receptor in POAG but our findings suggest a role for serum protein levels and support a causal role for this molecule in POAG pathophysiology.

Family history of Alzheimer’s disease and a diagnosis of squamous cell lung cancer were both found to have relationships with POAG within this study. We treat these results with a high degree of caution. The former is likely a product of violation of key assumptions of MR, whilst the latter is at high risk of survival bias. As such, whilst it is not possible to comment with certainty, we believe these results to be a consequence of bias. It is for instance understood that MR in elderly populations may produce results distorted by bias (as the analysis surrounds a nonrandom subset of the population who have survived long enough to develop a given disease) ^9,21^. With that in mind, it is evident that traits with a strong effect on survival are most likely to be biased and as such should be treated with a greater degree of caution. Additionally, recipients of care under urology were found to be more likely to develop POAG; this represents a broad composite trait inclusive of several confounding medical conditions, medications and other phenotypes and as such this result is not interpretable.

Our study has several limitations. First, we applied very strict criteria with respect to which traits we considered notable as we wanted to focus on the most robust results (which could then be explored in detail). As a result, our phenome-wide screen may have failed to identify true causal relationships for several traits (due to type 2 error). MR Egger is particularly underpowered relative to other MR tests and, whilst the inclusion of this method is likely to reduce the number of false positive findings, it may also lead to overlooking important traits. For instance, two potentially interesting proteins with roles in the trabecular meshwork (Serum inter-alpha-trypsin inhibitor heavy chain H and 72 kDa inositol polyphosphate 5-phosphatase) are significant in all measures except for MR Egger and could represent novel glaucoma-related molecules. This limitation applies to the other MR methods that we used, and a table presenting all results which were significant in FDR corrected IVW can be found in supplementary table 1.2 for readers to explore and research further. Second, as with all MR analyses, our results are vulnerable to violations of the assumptions of MR (including horizontal pleiotropy and unmeasured confounding) and, as such, no MR result in isolation can be categorically regarded to prove causation. Finally, in order to avoid bias due to population structure, the present analysis utilised data derived from populations of European ancestries only. This increases the risk of the results not being generalisable across populations.

In conclusion, this study provides support for a role for intraocular pressure and diabetes in POAG, suggests a protective effect for adiposity and highlights potential causal roles for serum cadherin 5 and angiopoietin-1 receptor. Future research on the effects of waist circumference, serum cadherin 5 and serum angiopoietin-1 receptor on POAG progression will provide key insights with a view to developing lifestyle modification advice and drug targets. Further, we suggest study of the local ocular effects of cadherin 5 and angiopoietin-1 receptor in order to glean mechanistic insights into the effects that topical interventions might have.

## METHODS

### Data sources

#### Exposure data

In this study we performed a phenome-wide screen to make causal inferences on the role of an extensive range of traits on POAG. To achieve this, we used both published and unpublished GWAS data from the MRC IEU open GWAS database (available online via https://gwas.mrcieu.ac.uk/); these were accessible via the TwoSampleMR programme in R ^37^. We included all ‘European GWAS’ within the database but removed imaging phenotypes and expression quantitative trait loci (eQTL) data due to difficulty in biological interpretation. ‘Finn’ trait GWAS data (derived from the FinnGen project) were excluded due to sample overlap. As it was not possible to manually inspect the degree of population overlap for all traits prior to conducting the analysis, the degree of overlap for all significant traits was inspected after the analysis.

#### Outcome data

The Gharahkhani *et al*. GWAS for POAG was utilised to provide the outcome data. Data were obtained from individuals of European ancestries only (16,677 POAG cases and 199,580 controls). Full details regarding the diagnosis of POAG and data sources contributing to the meta-analysis are detailed in the original paper ^7^. Where an exposure study was indexed in the MRC Integrative Epidemiology Unit (IEU) database with the prefix “ukb” or “met-d” (those which are explicitly 100% UK Biobank subjects) we utilised a version of the Gharahkhani *et al*. GWAS in which UK Biobank subjects were omitted (n= 15,229 POAG cases and 177,473 controls) ^38^. For all other studies, we manually inspected the composite populations if traits were identified as significant in order to ensure there was not significant sample overlap which might increase the risk of bias.

### Instrument selection

In this study, we used a statistically-driven approach to instrumental variable selection. Briefly, an arbitrary p value threshold was set for the identification of appropriate SNVs that could be used as instrumental variables (referred to thereafter as instruments). A conventional value for selection of instruments is p=5E-08, but this can, in some cases, be problematic. For example, when the number of instruments exceeding this threshold is small, the analysis can be underpowered or, in certain cases of unbiased screens, the results can be inflated^39^. With this in mind, we have set the p value for instrument selection for each trait to the level where >5 instruments are available for each analysis. More specifically, for each trait, the analysis would first be conducted with a p value for inclusion of 5E-8, and this would be sequentially increased by a factor of 10 each time until >5 eligible instruments are identified to a predefined maximum of p=5E-04. Ultimately, all significant traits were generated with instruments of p<5E-06.

### Proxies

Where an exposure instrument was not present in the outcome dataset we sought to identify a suitable proxy ^40^. This was achieved using the ensembl server with a linkage disequilibrium R^2^ value of 0.9 or above for identification of SNVs ^37,41^.

### Clumping

SNVs were clumped using a linkage disequilibrium R^2^ value of 0.001 and a genetic distance cut-off of 10,000 kilo-bases.

### Harmonisation

The effects of instruments on outcomes and exposures were harmonised in order to ensure that the beta values (i.e. the regression analysis estimates of effect size) were signed with respect to the same alleles ^40^. For palindromic alleles (i.e. alleles that are the same on the forward as on the reverse strand), those with minor allele frequency (MAF) > 0.42 were omitted from the analysis in order to reduce the risk of errors due to strand issues.

### Removal of pleiotropic genetic variants and outliers

In addition to using a range of MR methods and quality control measures (as detailed below), we endeavoured to remove pleiotropic instruments and outliers from the analysis. We operated this using ‘Steiger filtering’, a statistical approach in which instruments which were more significant in terms of p values for the outcome than the exposure were removed ^42^. Radial MR, an approach that detects and removes or downweights outlying instruments was also utilised for outlier identification ^43^.

### Causal inference

MR estimation was primarily performed using a multiplicative random effects (MRE) inverse variance weighted (IVW) analysis. MRE IVW was selected over a fixed effects (FE) approach as it allows inclusion of heterogeneous instruments, certain to occur within the breadth of this screen ^44^. MR relies on the selected instruments satisfying three core assumptions, these being that: (1) the instrumental SNVs are robustly associated with the exposure; (2) there must be no confounders of the SNV’s and the outcome; (3) the SNVs are associated with the disease risk outcome only through the exposure of interest ^9^. A range of methods were utilised to ensure the accuracy of the results and to cover for a range of violations of the aforementioned MR assumptions ^44^. The following approaches were selected: weighted median ^45^, MR Egger ^46^, weighted mode ^47^ and radial MR with modified second order weights ^43^.

### Further quality control

Instrument strength was determined using the F statistic (which tests the association between the instruments and the exposure) ^48^. F statistics were calculated against the final included set of instruments. Instrumental F statistics were calculated as ((N-1-k)/k)/(R^2^/(1-R^2^) where N = sample size, k = the number of instruments and R^2^ was the proportion of variance explained by the instrument. A mean F statistic >10 was considered sufficiently strong.

The Cochran’s Q test was performed for each analysis. Cochran’s Q is a measure of heterogeneity among causal estimates and serves as an indicator of the presence of horizontal pleiotropy (which occurs when an instrument exhibits effects on the outcome through pathways other than the exposure) ^49^. It is noted that a heterogeneous instrument is not necessarily invalid, but rather calls for a primary assessment with an MRE IVW rather than an FE approach; this has been conducted as standard throughout our analysis.

The MR Egger intercept test was used to detect horizontal pleiotropy. When this occurs, the MR Egger regression is robust to horizontal pleiotropy, under the assumption that that pleiotropy is uncorrelated with the association between the SNV and the exposure ^50^.

The I^2^ statistic was calculated as a measure of heterogeneity between variant specific causal estimates. An I^2^ <0.9 indicates that MR Egger is more likely to be biased towards the null through violation of the `NO Measurement Error’ (NOME) assumption ^51^.

Leave one out cross-validation was performed for every analysis to determine if any particular SNV was driving the significance of the causal estimates.

### Management of duplicate traits/GWAS

As the GWAS database used contains multiple different GWAS studies for certain traits, some exposures were analysed on multiple occasions. Where this occurred, we considered the largest sample size study to be the primary analysis. Where there were duplicate studies in the same population, we used the study with the largest F statistic. Where a binary trait was identified to have a causal relationship with POAG, we additionally ensured that the data utilised were the one that had the largest number of cases for the condition of interest.

### Identification of significant results

Before considering an MR result to be significant, the results of a range of causal inference and quality control tests should be taken into account. Notably, it is not necessary for a study to find significance in all measures to support a causal relationship. MR is a low power study type and, as such, an overly conservative approach to multiple testing can be excessive ^44^. However, in the context of the present study we have chosen to explore and discuss the results of our POAG phenome-wide screen only if they remained: significant after FDR correction in the MRE IVW; nominally significant in weighted mode, MR Egger and weighted median; and nominally significant throughout the leave one out analysis (MRE IVW) ^52^. This approach was selected as a large number of phenotypes were considered and because we wanted to focus on high-confidence signals to ensure reproducibility. Importantly, we expect our approach to overlook some important signals given the low power of our methodology. All our results are presented in the supplementary material and traits significant according to their FDR corrected p value in the IVW analysis represent phenotypes that may be of interest for further research and follow-up outside of this study.

### Presentation of effect sizes

Effect sizes are presented to enable readers to appraise the impact of putative risk factors. In univariable MR, these are shown as odds ratio (OR) per 1 standard deviation (SD) change in the exposure for continuous traits, and as beta values for binary exposure traits. This approach was chosen as odds ratios are generally uninformative for binary exposure traits ^53^.

### Rare variant burden testing

For significant protein phenotypes, we performed an orthogonal analysis using a gene-based rare variant burden test to aid functional interpretation. We utilised the AstraZeneca PheWas portal which includes whole genome sequencing data from the UK Biobank ^17^. Rare variants were defined as those having minor allele frequency less than 0.001 within both the UK Biobank and the Genome Aggregation Database (gnomAD) ^54^. A variant was assigned as deleterious if it was associated with a change in the amino-acid sequence and reached a REVEL score >0.25 ^55^. Variants that met these criteria were collapsed into a single two-sided Fisher’s exact test to assess the relative frequency in individuals with POAG and in unaffected controls. Our analysis included 3,088 POAG cases and 339,310 controls. A rare variant burden reaching nominal significance was considered significantly associated with POAG.

### Software

R version 4.1.0.

TwoSampleMR version 0.5.6.

RadialMR version 1.0.

## Data Availability

All data produced in this study are available in the supplementary content.

## Ethical approval

This study utilised publicly available data and no additional ethical approval was required.

## Data availability

All data associated with this study are published in the supplementary material.

## Author contributions

THJ, GCB, JCK, ES and PIS were involved in study design, data analysis, data interpretation, and manuscript write up. HG, JY and ZG were involved in data interpretation and manuscript write up. All authors approved the final manuscript.

## Acknowledgements

We acknowledge the following sources of funding: the Wellcome Trust (224643/Z/21/Z, Clinical Research Career Development Fellowship to P.I.S.; 200990/Z/16/Z, Transforming Genetic Medicine Initiative to G.C.B.); the UK National Institute for Health Research (NIHR) Clinical Lecturer Programme (CL-2017-06-001 to P.I.S.); the UK NIHR Academic Clinical Fellowship Programme (T.H.J); Retina UK and Fight for Sight (GR586, RP Genome Project—UK Inherited Retinal Disease Consortium to G.C.B.); the University of Manchester’s Wellcome Institutional Strategic Support Fund (Wellcome ISSF) grant (204796/Z/16/Z).

